# Diabetes and all-cause mortality among middle-aged and older adults in China, England, Mexico, rural South Africa, and the United States: A population-based study of longitudinal aging cohorts

**DOI:** 10.1101/2024.10.09.24315174

**Authors:** Hunter Green, Yuan S. Zhang, Chihua Li, Paola Zaninotto, Kenneth M. Langa, Jinkook Lee, Jennifer Manne-Goehler, David Flood

## Abstract

**Objective:** There is a need for comparable worldwide data on the impact of diabetes on mortality. This study assessed diabetes and all-cause mortality among middle-aged and older adults in five countries.

**Research Design and Methods:** We analyzed adults aged 51 years or older followed between 2010 and 2020 from population-based cohorts in China, England, Mexico, rural South Africa, and the United States. Diabetes was defined by self-report or an elevated diabetes blood-based biomarker meeting the clinical criteria for diabetes. All-cause mortality was assessed through linkages or informant interviews. We used Poisson regression models to estimate mortality rate ratios and mortality rate differences, comparing people with diabetes to those without diabetes. Models were adjusted for age, gender, education, smoking status, body mass index, and economic status.

**Results:** We included 29,397 individuals, of whom 4,916 (16.7%) died during the study period. The median follow-up time ranged from 4.6 years in South Africa to 8.3 years in China. The adjusted all-cause mortality rate ratios for people with diabetes versus those without diabetes ranged from 1.53 (95% CI: 1.39-1.68) in the United States to 2.02 (95% CI: 1.34-3.06) in Mexico. The adjusted mortality rate differences (per 1,000 person-years) for people with diabetes versus those without diabetes ranged from 11.9 (95% CI: 4.8-18.9) in England to 24.6 (95% CI: 12.2-37.0) in South Africa.

**Conclusions:** Diabetes was associated with increased all-cause mortality in population-based cohorts across five diverse countries. There is an urgent need to implement clinical and public health interventions to improve diabetes outcomes globally.

**ARTICLE HIGHLIGHTS:** *Why did we undertake this study?:* We aimed to address the need for comparable estimates of all-cause mortality among people with diabetes in diverse global settings.

*What is the specific question(s) we wanted to answer?:* How does diabetes impact all-cause mortality among middle-aged and older adults (aged 51 years or greater) in China, England, Mexico, rural South Africa, and the United States?

*What did we find?:* Middle-aged and older adults with diabetes had higher all-cause mortality than people without diabetes in all countries. Relative mortality differences ranged from mortality rate ratios of 1.53 in the United States to 2.02 in Mexico. Absolute mortality differences ranged from mortality rate differences (per 1,000 person-years) of 11.9 in England to 24.6 in South Africa.

*What are the implications of our findings?:* There is an urgent need to implement clinical and public health interventions to improve diabetes outcomes globally.

## INTRODUCTION

More than half a billion people worldwide are living with diabetes.^1,2^ By 2050, this number will increase to 1.2 billion people.^1^ Given the epidemiology of diabetes, it is crucial to understand how it impacts long-term health outcomes such as mortality in diverse populations worldwide. All-cause mortality among people with diabetes at the population level is a key metric in the World Health Organization (WHO) global diabetes monitoring framework.^3^ The WHO recommends monitoring diabetes mortality because it is inherently significant to patients and policymakers, modifiable through evidence-based interventions, and amenable to standardized assessment methods.^3^

While diabetes has long been associated with increased mortality in high-income countries,^4–6^ contemporary and cross-national estimates of this association have been limited by several factors. First, there is a paucity of data on diabetes and mortality from low- and middle-income countries where most people with diabetes live, and this is especially true for middle-aged or older adults who are often understudied in these settings.^7^ Second, temporal declines in all-cause mortality in high-income countries have been observed in recent decades, so updated data are needed.^8^ Third, population data on diabetes and mortality are often not comparable across settings due to differences in sample selection, case definitions, and mortality ascertainment.^7^ These limitations pose challenges for accurately assessing the global burden of diabetes and monitoring diabetes policy responses.

To address these gaps, this study aimed to evaluate the association between diabetes and all-cause mortality among middle-aged and older adults with diabetes using recent data from comparable population-based aging cohorts in five diverse countries.

## RESEARCH DESIGN AND METHODS

### Study design and sample

We conducted a longitudinal analysis of population-based aging cohorts in five countries: China (China Health and Retirement Longitudinal Study [CHARLS]),^9^ England (English Longitudinal Study of Ageing [ELSA]),^10^ Mexico (Mexican Health and Aging Study [MHAS]),^11^ rural South Africa (Health and Aging in Africa: A Longitudinal Study of an INDEPTH Community in South Africa [HAALSI]),^12^ and the United States (Health and Retirement Study [HRS]).^13^ These cohorts are part of the HRS International Family of Studies, a network of longitudinal aging studies with similar sampling designs, eligibility, and assessment methods.^14^ The cohort inclusion criteria for this analysis were (1) availability of baseline and follow-up data from 2010 to 2020 and (2) collection of a blood-based diabetes biomarker at the baseline wave during this period. We chose 2010 to 2020 as our period of interest to maximize comparability between cohorts. With one exception, the included cohorts were nationally representative of each country’s middle-aged and older population. The exception was the South Africa cohort, which was representative of rural communities in sub-Saharan Africa. See Appendix 1 for details on the years of data collection and censoring by cohort.

Due to minor differences in the lower end of age eligibility between cohorts, we excluded individuals younger than 51 at baseline to ensure comparability. We also excluded respondents without follow-up information, with no available blood-based diabetes biomarker, or with missing data on prior diabetes diagnosis, gender, education, economic status, smoking status, BMI, or survey weights. In the South Africa cohort, we excluded individuals with missing human immunodeficiency virus (HIV) status. Appendix 2 shows participant flow diagrams for each cohort including numbers lost to follow-up.

### Definition of diabetes

We defined diabetes as either (1) a history of self-reported diagnosis by a physician or health care worker or (2) an elevated blood-based biomarker meeting clinical criteria for diabetes.^15,16^ We used HbA1c ≥6.5% (48 mmol/mol) as the biomarker threshold in all countries except China and South Africa where we used fasting blood glucose ≥126 mg/dL (7.0 mmol/L) or random blood glucose ≥200 mg/dL (11.1 mmol/L). In China, plasma glucose was assessed using an enzymatic colorimetric test (92% of individuals were fasting). In the South African cohort, capillary glucose was assessed using a point-of-care analyzer (24% of individuals were fasting). In England, HbA1c was assessed using venous blood samples. In Mexico, HbA1c was assessed using a point-of-care analyzer certified by the National Glycohemoglobin Standardization Program.^17^ In the United States, HbA1c was assessed using dried blood spots converted to whole blood equivalent values.^18^ Relevant question text and biomarker details are provided in Appendix 3-4.

### Mortality ascertainment

All-cause mortality was captured in England by linking to the National Health Service Central Register (latest available data from April 2018). In other cohorts, all-cause mortality was captured during interviews with respondents’ spouses or other informants. In all cohorts, the month and year of death were available. If the date of death was unknown, it was estimated as the midpoint between waves in which an individual was known to be alive and had died. We measured survival time in years from the baseline interview as defined in this study to death, loss to follow-up, or the end of the follow-up period (May 2018 in England and December 2019 in the other countries), whichever came first.

### Statistical analysis

Analyses were conducted within each cohort and accounted for survey weights and sampling design when available. We first calculated the overall and age-stratified diabetes prevalence at baseline. In calculating overall prevalence, we age-standardized to the WHO standard population. We then used Poisson regression with an offset for log-transformed person-years and robust standard errors to estimate differences in mortality rates among people with diabetes versus those without diabetes. Poisson models give similar results to Cox models when there are shorter follow-up intervals and have the advantage of directly estimating event rates.^19,20^ Both relative (mortality rate ratios) and absolute (mortality rate and mortality rate differences) measures are reported. Mortality rates and mortality rate differences are presented as the number of deaths per 1,000 person-years.

We used prior evidence to develop a directed acyclic graph showing our conceptual model of the relationship between diabetes and mortality (Appendix 5).^21^ We adjusted for baseline covariates, including age (51-59 years, 60-69 years, and ≥70 years), gender (women vs. men), education (less than upper secondary, upper secondary and vocational, and tertiary), smoking status (current vs. not current smoker), BMI categories (underweight: BMI < 18.5 kg/m^2^; normal weight: 18.5-24.9 kg/m^2^; overweight: 25.0-29.9 kg/m^2^; obese: ≥ 30.0 kg/m^2^), and economic status (tertiles). Economic status was defined as the annual income of the individual and their co-residing spouse or dependent children in high-income countries (England and the United States) and the annual household per-capita consumption in upper-middle-income countries (China, Mexico, and South Africa). Per-capita consumption is the preferred measure of living standard derived from surveys in developing countries.^22^ In the South African cohort, we also adjusted for HIV status, given the high prevalence (23%) and known mortality association in this population.^23^ Models were run in the overall sample and by gender. We estimated differences in mortality rates among people with no diabetes, diagnosed diabetes, and undiagnosed diabetes. Analyses were performed using Stata version 18.0.

### Sensitivity analyses

We conducted three sensitivity analyses. First, we assessed the consistency of our findings using Cox proportional hazards regression models. Second, we estimated the association between diabetes and mortality using a slightly more restrictive epidemiological diabetes definition of either (1) the self-reported use of a glucose-lowering medication or (2) an elevated biomarker meeting clinical criteria for diabetes.^24,25^ Finally, we performed an analysis without the adjustment for BMI categories given the potentially bidirectional relationship between diabetes and BMI.

### Data availability and ethics

This study was deemed exempt from institutional ethics approval at the University of Michigan (HUM00256096). Data included in this study are publicly available for all cohorts except for mortality data for ELSA. Details on accessing data can be found at the Gateway to Global Aging Data website (https://g2aging.org/).

## RESULTS

### Survey and respondent characteristics

Table 1 presents survey and respondent characteristics for the five cohorts. The final sample included 6,251 individuals in China, 4,819 in England, 1,717 in Mexico, 3,411 in South Africa, and 13,199 in the United States. Of the 29,397 total individuals, 4,916 (16.7%) died during the study period. The median follow-up time ranged from 4.6 (interquartile range [IQR]: 4.4-4.8) years in South Africa to 8.3 (IQR: 8.2-8.4) years in China. There were 191,782 total person-years of follow-up in the cohorts (China: 48,122 person-years; England: 24,536 person-years; Mexico: 11,192 person-years; South Africa: 14,722 person-years; and United States: 93,210 person-years).

**Table 1:** Survey and respondent characteristics.

|  | **China**<br>**(CHARLS)** | **England**<br>**(ELSA)** | **Mexico**<br>**(MHAS)** | **South Africa**<br>**(HAALSI)** | **United States**<br>**(HRS)** |
| --- | --- | --- | --- | --- | --- |
| **Survey characteristics** |  |  |  |  |  |
| Years of data collection (baseline to endline) | 2011 to 2019 | 2012 to 2018 | 2012 to 2019 | 2014 to 2019 | 2010 to 2019 |
| Sample size, n | 6,251 | 4,819 | 1,717 | 3,411 | 13,199 |
| Deaths, n | 968 | 400 | 206 | 510 | 2,832 |
| Follow-up time (years), median (IQR) | 8.3 (8.2-8.4) | 5.5 (5.2-5.7) | 7.1 (7.1-7.2) | 4.6 (4.4-4.8) | 7.5 (6.2-9.0) |
| **Respondent characteristics** |  |  |  |  |  |
| Age (years), median (IQR) | 61 (56-68) | 64 (57-72) | 63 (56-70) | 65 (57-73) | 63 (57-72) |
| Women, % (95% CI) | 50.2 (47.8-52.6) | 50.9 (49.4-52.3) | 55.2 (50.8-59.6) | 53.7 (52.0-55.4) | 54.1 (53.3-54.9) |
| Education, % (95% CI) |  |  |  |  |  |
| Less than upper secondary | 89.9 (88.2-91.4) | 30.8 (29.3-32.5) | 88.8 (85.9-91.2) | 93.3 (92.4-94.1) | 13.9 (12.7-15.4) |
| Upper secondary and vocational | 8.2 (7.3-9.2) | 50.7 (49.0-52.4) | 3.4 (2.1-5.6) | 4.3 (3.7-5.0) | 58.7 (56.9-60.4) |
| Tertiary | 1.9 (0.9-3.9) | 18.4 (17.1-19.9) | 7.7 (5.9-10.1) | 2.4 (1.9-2.9) | 27.4 (25.5-29.4) |
| Current smoker, % (95% CI) | 31.0 (28.6-33.6) | 13.7 (12.5-15.0) | 16.9 (12.9-21.8) | 8.4 (7.5-9.3) | 14.9 (13.8-16.0) |
| BMI, % (95% CI) |  |  |  |  |  |
| <18.5 (Underweight) | 7.5 (6.7-8.3) | 0.9 (0.6-1.3) | 0.9 (0.4-1.8) | 5.6 (4.9-6.5) | 1.0 (0.8-1.3) |
| 18.5-24.9 (Normal) | 61.0 (58.6-63.4) | 26.4 (25.0-28.0) | 26.0 (21.8-30.7) | 36.5 (34.9-38.1) | 20.8 (20.0-21.7) |
| 25.0-29.9 (Overweight) | 27.0 (24.6-29.5) | 41.8 (40.2-43.5) | 38.0 (34.0-42.2) | 28.3 (26.9-29.9) | 34.8 (33.8-35.8) |
| ≥30 (Obese) | 4.6 (3.9-5.3) | 30.8 (29.3-32.4) | 35.1 (31.0-39.5) | 29.6 (28.0-31.1) | 43.3 (42.3-44.4) |
| Diabetes (diagnosed and undiagnosed), % (95% CI)* | 15.7 (14.3-17.2) | 11.7 (10.6-13.0) | 37.4 (33.4-41.5) | 12.1 (11.1-13.3) | 21.8 (20.8-22.8) |
| Diagnosed among all with diabetes, % (95% CI)* | 46.4 (42.1-50.7) | 76.1 (69.3-81.7) | 53.6 (47.2-59.9) | 57.8 (52.5-62.9) | 86.1 (84.1-87.9) |

\*Values are age-standardized to the WHO standard population among adults aged 50 years and older. CHARLS=China Health and Retirement Longitudinal Study. ELSA=English Longitudinal Study of Ageing. HAALSI=Health and Aging in Africa: A Longitudinal Study of an INDEPTH Community in South Africa. HRS=Health and Retirement Study. IQR=Interquartile range. MHAS=Mexican Health and Aging Study.

There was considerable cross-country variation in some respondent characteristics, as illustrated in Table 1. For example, while nine-tenths of individuals in China (89.9% [95% CI: 88.2-91.4]), Mexico (88.8% [95% CI: 85.9-91.2]), and South Africa (93.3% [95% CI: 92.4-94.1]) had less than an upper secondary education, most individuals in England (69.2% [95% CI: 67.5-70.8]) and the United States (86.1% [95% CI: 84.7-87.4]) had an upper secondary education or greater. Current smoking ranged from 8.4% (95% CI: 7.5-9.3) in South Africa to 31.0% (95% CI: 28.6-33.6) in China. The prevalence of individuals who were obese ranged from 4.5% (95% CI: 3.9-5.3) in China to 43.3% (95% CI: 42.3-44.4) in the United States.

### Diabetes prevalence

The age-standardized prevalence of diabetes was highest in Mexico (37.4% [95% CI: 33.4-41.5], followed by the United States (21.8% [95% CI: 20.8-22.8]), China (15.7% [95% CI: 14.3-17.2]), South Africa (12.1% [95% CI: 11.1-13.3]), and England (11.7% [95% CI: 10.6-13.0]). Figure 1 shows the age-specific prevalence of diabetes by cohort at baseline. Among individuals with diabetes, the age-standardized proportion of those with diabetes who reported a prior diabetes diagnosis ranged from 46.4% (95% CI: 42.1-50.7) in China to 86.1% (95% CI: 84.1-87.9) in the United States (Table 1).

**Figure 1:**
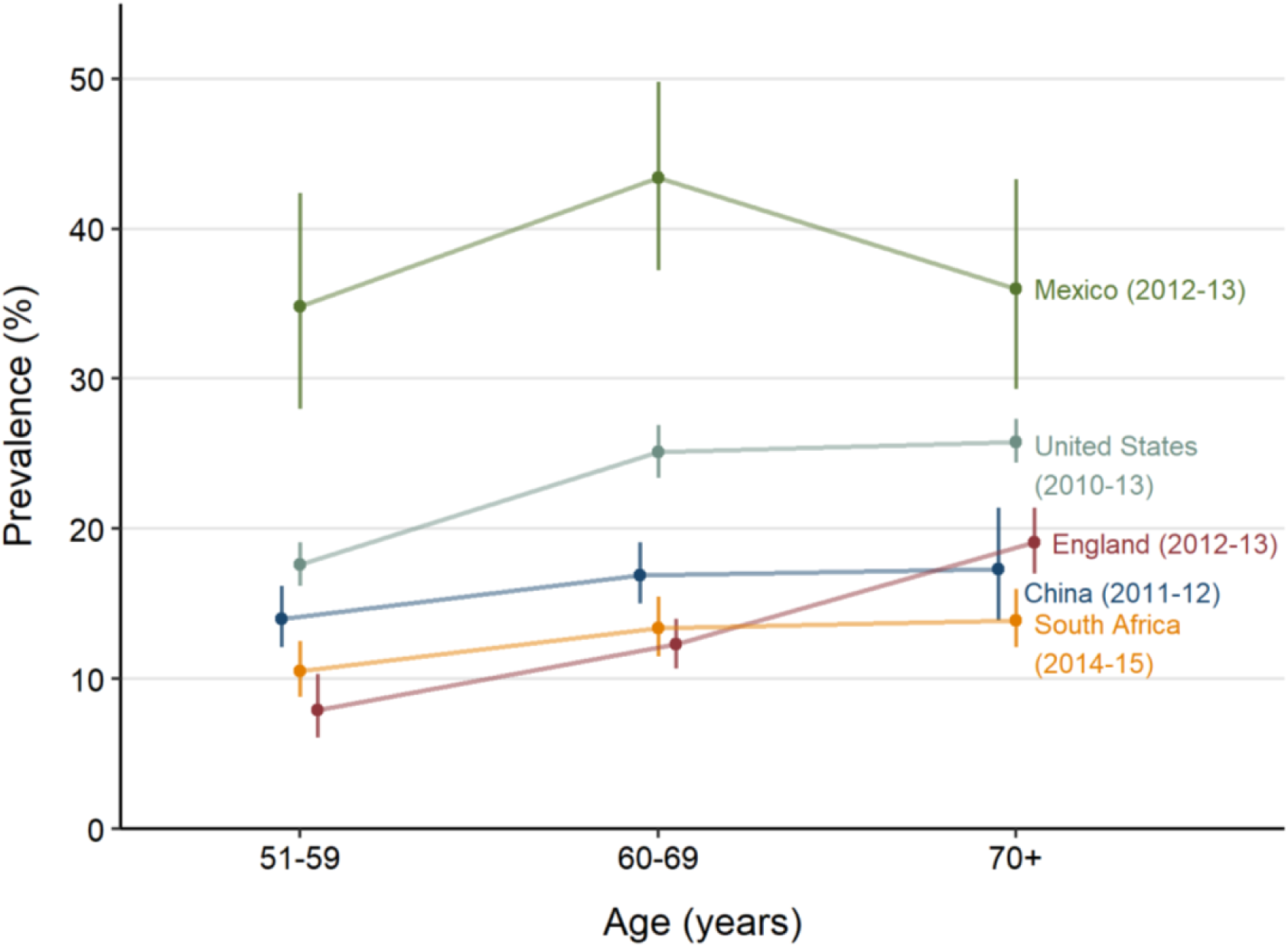
Age-specific prevalence of diabetes by cohort. Diabetes was defined among individuals self-reporting a previous diabetes diagnosis or those with an elevated biomarker (hemoglobin A1c ≥6.5% [48 mmol/mol], fasting plasma glucose ≥126 mg/dL [7.0 mmol/L], or random capillary glucose ≥200 mg/dL [11.1 mmol/L]). The vertical error bars represent 95% CIs.

### Mortality rates

Adjusted all-cause mortality rates (per 1,000 person-years) are presented in Figure 2 and Appendix 6. In each cohort, mortality rates were higher among people with diabetes than those without diabetes. Across the cohorts, mortality rates among people with diabetes were highest in South Africa (57.5 [95% CI: 45.5-69.5]), followed by the United States (39.2 [95% CI: 36.1-42.4]), China (95% CI: 35.5 [95% CI: 28.6-42.4]), England (28.8 [95% CI: 22.1-35.6]), and Mexico (29.0 [95% CI: 19.0-39.0]).

**Figure 2:**
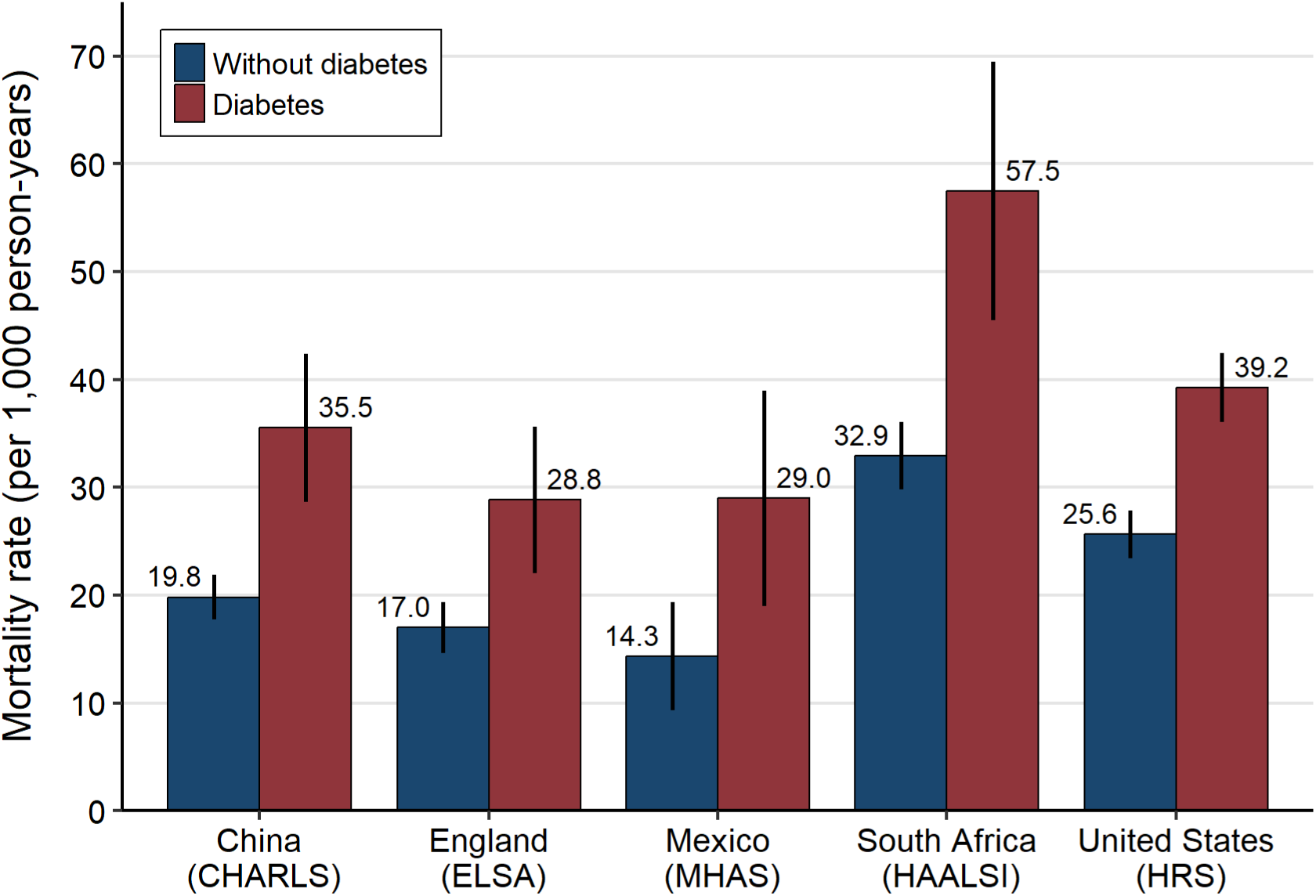
Adjusted all-cause mortality rates by cohort. Mortality rates are presented as the number of deaths per 1,000 person-years. The vertical error bars represent 95% CIs. Estimates were derived using Poisson regression models with an offset for log-transformed person-years and robust standard errors and adjusted for age, gender, education, smoking status, body mass index, and economic status. Models in South Africa also adjusted for HIV status. CHARLS=China Health and Retirement Longitudinal Study. ELSA=English Longitudinal Study of Ageing. HAALSI=Health and Aging in Africa: A Longitudinal Study of an INDEPTH Community in South Africa. HRS=Health and Retirement Study. MHAS=Mexican Health and Aging Study.

### Mortality rate ratios and mortality rate differences

The adjusted overall all-cause mortality rate ratios for people with diabetes versus those without diabetes ranged from 1.53 (95% CI: 1.39-1.68) in the United States to 2.02 (95% CI: 1.34-3.06) in Mexico (Figure 3A). The adjusted mortality rate differences (per 1,000 person-years) for people with diabetes versus those without diabetes ranged from 11.9 (95% CI: 4.8-18.9) in England to 24.6 (95% CI: 12.2-37.0) in South Africa. No significant differences were observed in adjusted mortality rate ratios or adjusted mortality rate differences by sex in cohorts. Mortality rate ratios appeared to decrease in older age groups in the cohorts from England and the United States (Appendix 7).

**Figure 3:**
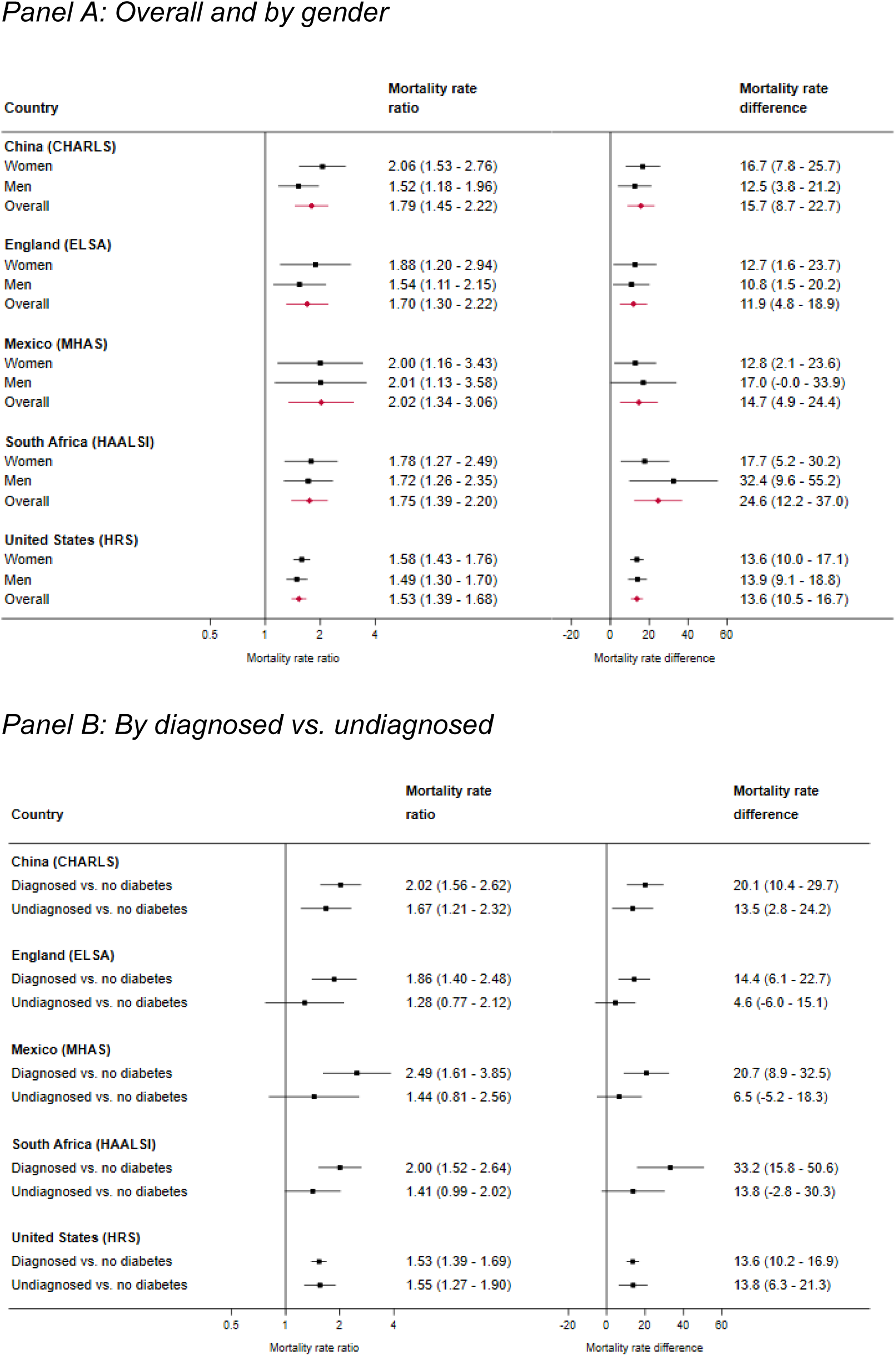
Adjusted all-cause mortality rate ratios and mortality rate differences. Mortality rate differences are presented as the number of deaths per 1,000 person-years. The horizontal error bars represent 95% CIs. Estimates were derived using Poisson regression models with an offset for log-transformed person-years and robust standard errors and adjusted for age, gender, education, smoking status, body mass index, and economic status. Models in South Africa also adjusted for HIV status. CHARLS=China Health and Retirement Longitudinal Study. ELSA=English Longitudinal Study of Ageing. HAALSI=Health and Aging in Africa: A Longitudinal Study of an INDEPTH Community in South Africa. HRS=Health and Retirement Study. MHAS=Mexican Health and Aging Study.

Figure 3B shows results when the diabetes classification was separated by diagnosed or undiagnosed status, compared to no diabetes. In general, there appeared to be a tendency among people diagnosed with diabetes to have higher mortality than people with undiagnosed diabetes. However, these differences were statistically significant only in Mexico where people with diagnosed diabetes compared to undiagnosed diabetes had an adjusted mortality rate ratio of 1.95 (95% CI: 1.11-3.43), corresponding to an adjusted mortality rate difference of 22.6 (95% CI: −7.3-52.4) deaths per 1,000 person-years (Appendix 8).

### Sensitivity analyses

The results of the first sensitivity analysis using Cox proportional hazards regression models (Appendix 9) were generally consistent with the main results using Poisson regression models. In the second sensitivity analysis using the slightly more restrictive epidemiological diabetes definition of either the use of a glucose-lowering medication (in place of self-reported diagnosis) or an elevated biomarker, we observed a slightly higher point estimate for the adjusted mortality rate ratios in the China cohort (1.86 vs. 1.79) and slightly lower adjusted mortality rate ratios in the Mexico cohort (1.84 vs. 2.02; Appendix 10). The third sensitivity analysis removing adjustment for BMI had the effect of slightly attenuating the mortality rate ratios and mortality rate differences compared to the main analysis (Appendix 11).

## CONCLUSIONS

In this study of middle-aged and older adults followed between 2010 and 2020 from population-based cohorts in five diverse countries (four of which were nationally representative), we found that people with diabetes consistently had higher all-cause mortality than people without diabetes. Relative mortality differences were similar across cohorts, ranging from mortality rate ratios of 1.53 (95% CI: 1.39-1.68) in the United States to 2.02 (95% CI: 1.34-3.06) in Mexico. Absolute mortality differences had more variation across cohorts, ranging from mortality rate differences (per 1,000 person-years) of 11.9 (95% CI: 4.8-18.9) in England to 24.6 (95% CI: 12.2-37.0) in South Africa. These findings using recent and comparable data highlight the immense burden of diabetes around the world, particularly in low- and middle-income country settings such as rural South Africa and Mexico where the absolute mortality impact of diabetes appears greatest. These are also settings where diabetes care is thought to be least robust.^3,26,27^

Many prior studies assessing the association between diabetes and all-cause mortality have been conducted in high-income countries and among younger age groups.^3,7,8^ Large-scale meta-analyses in the last two decades have reported relative mortality differences among people with diabetes, as compared to those without diabetes, that are generally similar to findings in our study.^4,6,28,29^ However, these meta-analyses primarily included non-representative cohorts from high-income countries, limiting population inferences globally. A multi-country analysis from 1995 to 2016 in 16 countries provides updated evidence of a reduction in all-cause mortality among people with diagnosed diabetes, but data were only available from high-income countries.^8^ The Prospective Urban Rural Epidemiology study reported greater absolute mortality among people with diabetes in middle-income and low-income countries, as compared to people with diabetes in high-income countries.^5^ While studies on diabetes-related mortality previously have been performed in each of the countries included in our analysis,^30–36^ our study uniquely compares diabetes-related mortality in multiple countries using similar methods across the entire continuum of middle-aged and older adults. Individuals in this age range are sometimes excluded from population-based studies worldwide. However, they have the highest diabetes prevalence and require comprehensive clinical management to prevent diabetes complications.

An important secondary finding in our study was the tendency of higher mortality among people with diagnosed diabetes compared to undiagnosed diabetes. This finding was most marked in Mexico and South Africa. Many—though not all—prior high-quality population-based studies have reported similar findings.^5,30–32,37^ We hypothesize that the greater mortality among people with previously diagnosed compared to undiagnosed diabetes likely reflects a selection effect related to diabetes severity and/or diabetes duration. Diabetes patients with the highest disease severity or progression are most likely to experience symptoms, to seek a diagnosis in the health care system, and, despite obtaining a diagnosis, to die. This selection effect may be most salient in countries at lower income levels where the proportion of adults with diabetes who are diagnosed is as low as 20%, compared to 80% or greater in some high-income countries such as the United States.^3^

What are the policy implications emerging from this work? The higher absolute mortality rates in South Africa and Mexico suggest that people with diabetes in these countries experience challenges accessing quality diabetes care. There is an urgent need to scale up evidence-based interventions to manage diabetes and its associated cardiovascular risk factors, particularly in low- and middle-income countries where societies are aging, absolute diabetes mortality is highest, and the population with diabetes is rapidly growing.^2^ Evidence from Sweden shows that people with diabetes who are appropriately managed and achieve risk factor control have little or no excess mortality compared to those without diabetes.^38^ Yet only 10% of people with diabetes in low- and middle-income countries receive comprehensive diabetes management aligned with guidelines.^26^ In the coming decades, diabetes will cause a staggering degree of premature mortality unless health systems are strengthened to improve diabetes care.^1^ The WHO Global Diabetes Compact is a crucial international effort to stimulate improvements in equitable, affordable, and quality care for people with diabetes.^1,3^ A key pillar of these efforts is the inclusion of stakeholders from the public and private sectors, as well as individuals with lived experiences of diabetes.

Our study has several limitations. First, our analysis did not include people aged 50 years or younger. Younger populations with diabetes tend to have high diabetes mortality compared to young populations without diabetes.^30–32^ Our results should not be generalized to entire populations or young populations. Still, they can be generalized to populations aged 51 years or older, which represent approximately two-thirds of people with diabetes worldwide.^2^ Second, differences in the blood-based diabetes biomarkers collected in each cohort (e.g., glucose versus HbA1c) may contribute to slightly different phenotypes of individuals classified as having undiagnosed diabetes.^39,40^ This limitation could decrease the comparability of estimates across cohorts. Third, our study lacks data on cause-specific mortality, preventing us from distinguishing between microvascular and macrovascular patterns of death among individuals with diabetes. Fourth, the South African cohort was not nationally representative, though it is representative of a rural community like many others in sub-Saharan Africa. Finally, while this analysis used data from a geographically and economically diverse set of countries, the included cohorts may not fully represent populations with diabetes worldwide. In particular, none of the cohorts were drawn from low- or lower-middle-income countries. Estimating diabetes mortality in these settings is an important area of future research.

In summary, we observed that diabetes was consistently associated with increased all-cause mortality across five diverse settings, and absolute diabetes mortality was particularly high in low- and middle-income countries where systems of care for diabetes are known to be weaker. The findings reinforce the need to implement clinical and public health interventions to improve diabetes outcomes in countries worldwide.

## Funding and Assistance

The preparation of this paper was supported by the Gateway to Global Aging Data, which is funded by the U.S. National Institute on Aging (award R01AG030153). The English Longitudinal Study of Ageing is funded by the National Institute on Aging (award R01AG017644) and by a consortium of U.K. government departments (Department for Health and Social Care; Department for Transport; and Department for Work and Pensions, which is coordinated by the National Institute for Health Research [NIHR, Ref: 198-1074]). The Health and Retirement Study is funded by the U.S. National Institute on Aging (award U01AG009740) and the Social Security Administration, and performed at the Institute for Social Research, University of Michigan. The Health and Aging in Africa: A Longitudinal Study in South Africa study is funded by the U.S. National Institute on Aging (award 5P01AG041710). D.F. was supported by the U.S. National Heart, Lung, and Blood Institute (award K23HL161271), the Michigan Center for Diabetes Translational Research (award P30DK092926), the University of Michigan Claude D. Pepper Older Americans Independence Center (award 5P30AG024824), and the University of Michigan Caswell Diabetes Institute Clinical Translational Research Scholars Program. Y.S.Z was supported by the U.S. National Institute on Aging (award R00AG070274). JM-G is supported by the National Institute of Diabetes and Digestive and Kidney Diseases, National Institutes of Health (award number K23DK12516). The contents of this research are solely the responsibility of the authors and do not necessarily represent the official views of the National Institutes of Health.

## Author Contributions and Guarantor Statement

H.G., J.M.-G., and D.F. conceived the idea for this study. H.G. conducted the statistical analysis. H.G. and D.F. wrote the first draft of the manuscript with substantial revisions from J.M.-G. H.G. and P.Z. verified the underlying data. Y.S.Z. and P.Z. provided analytic support. All authors provided crucial input on multiple iterations of the manuscript. D.F. had full access to the data except the ELSA mortality data; due to privacy regulations, these data were restricted in access to P.Z. D.F. is the guarantor of this work and, as such, takes responsibility for the integrity of the data and the accuracy of the data analysis.

