## Supplementary appendix for "Diabetes and all-cause mortality among middle-aged and older adults in China, England, Mexico, rural South Africa, and the United States: A population-based study of longitudinal aging cohorts"

**Supplementary data**

**Appendix 1: Years of data collection and censoring by cohort**

| Study | Years of data collection and censoring |
| --- | --- |
| China (CHARLS) | Respondents were followed from the 2011-2012 wave to the 2020 wave. Respondents were right censored at their last interview date or December 2019, whichever came first. |
| England (ELSA) | Respondents were followed from the 2012-2013 wave to the 2018-2019 wave. Respondents were right censored at their last interview date or May 2018, whichever came first. |
| Mexico (MHAS) | Respondents were followed from the 2012-2013 wave to the 2021-2022 wave. Respondents were right censored at their last interview date or December 2019, whichever came first. |
| South Africa (HAALSI) | Respondents were followed from the 2014-2015 wave to the 2019-2022 wave. Respondents were right censored at their last interview date or December 2019, whichever came first. |
| United States (HRS) | Respondents were followed from the 2010-2011 wave or the 2012-2013 wave to the 2020-2021 wave. Respondents were right censored at their last interview date or December 2019, whichever came first. |

**Appendix 2: Analysis flow diagrams**

China Health and Retirement Longitudinal Study (CHARLS)

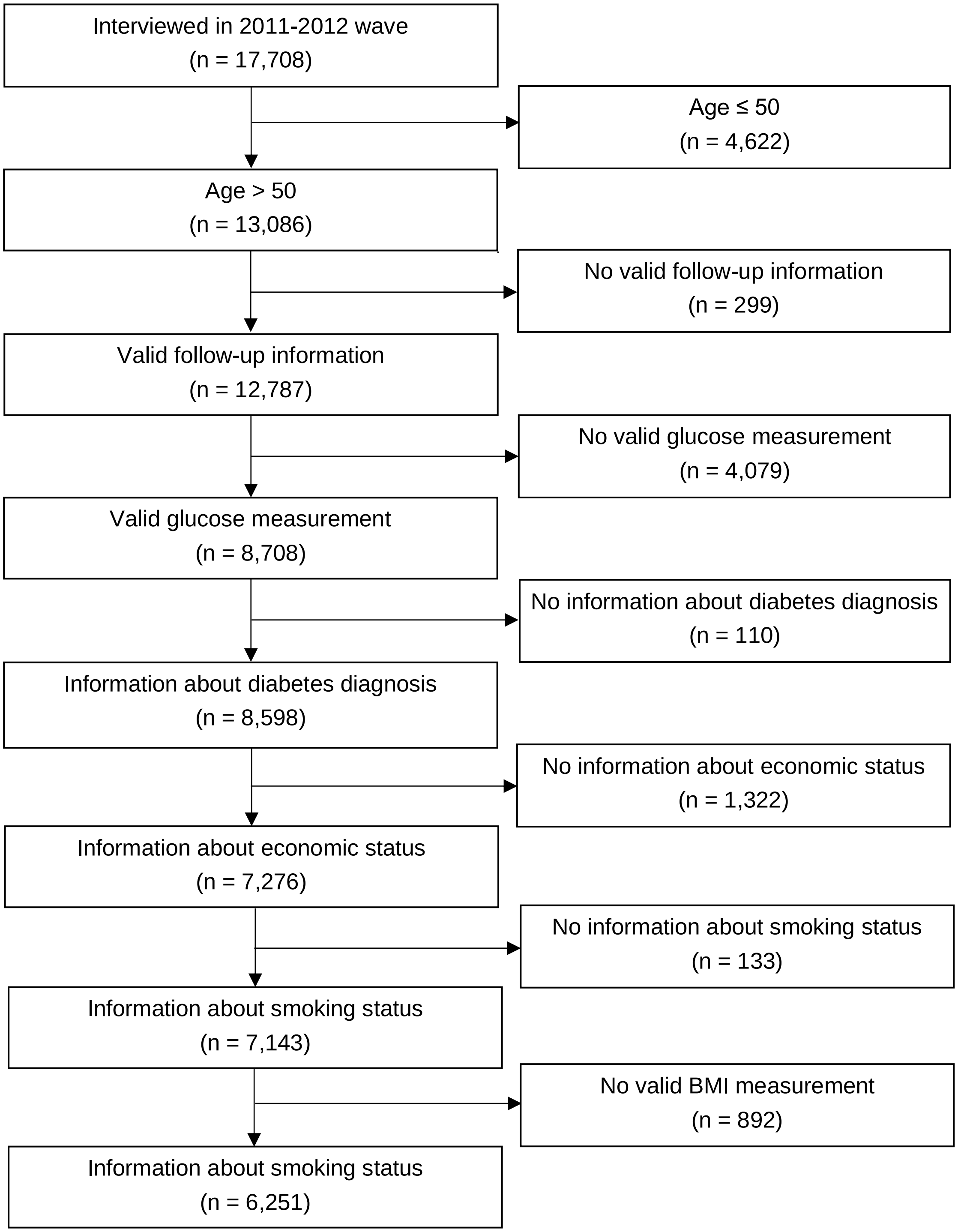

English Longitudinal Study of Ageing (ELSA)

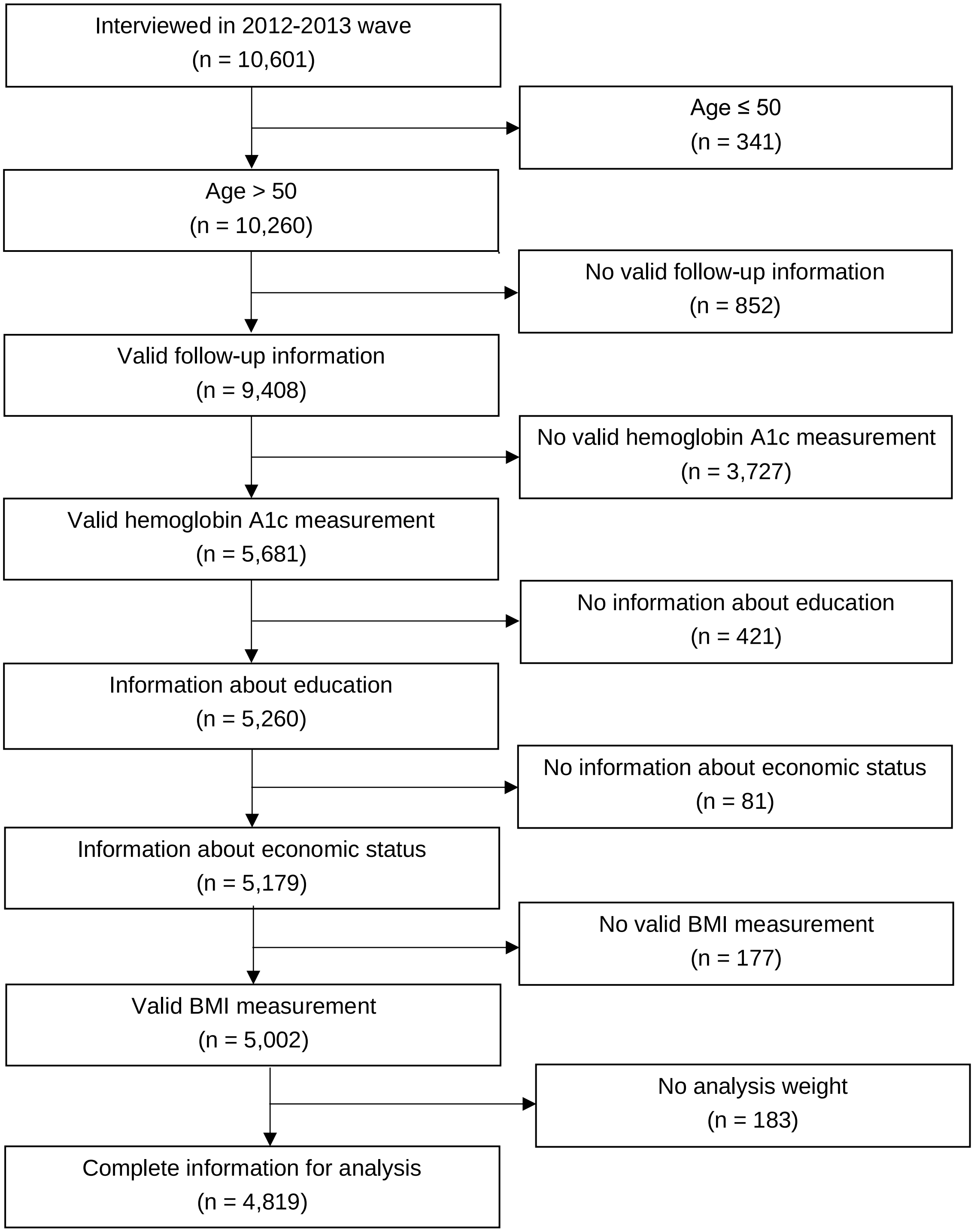

Mexican Health and Aging Study (MHAS)

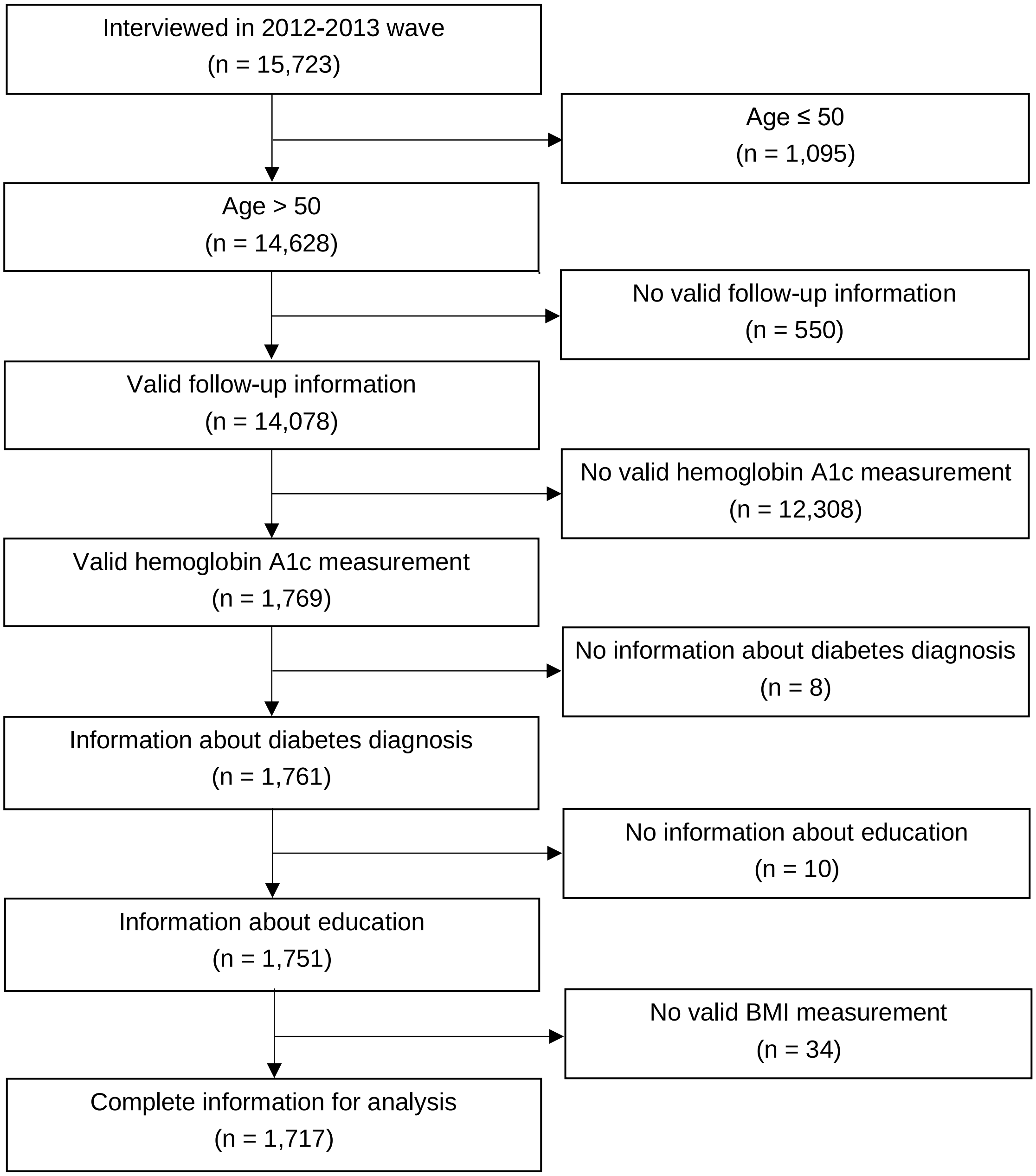

Health and Aging in Africa: A Longitudinal Study of an INDEPTH Community in South Africa (HAALSI)

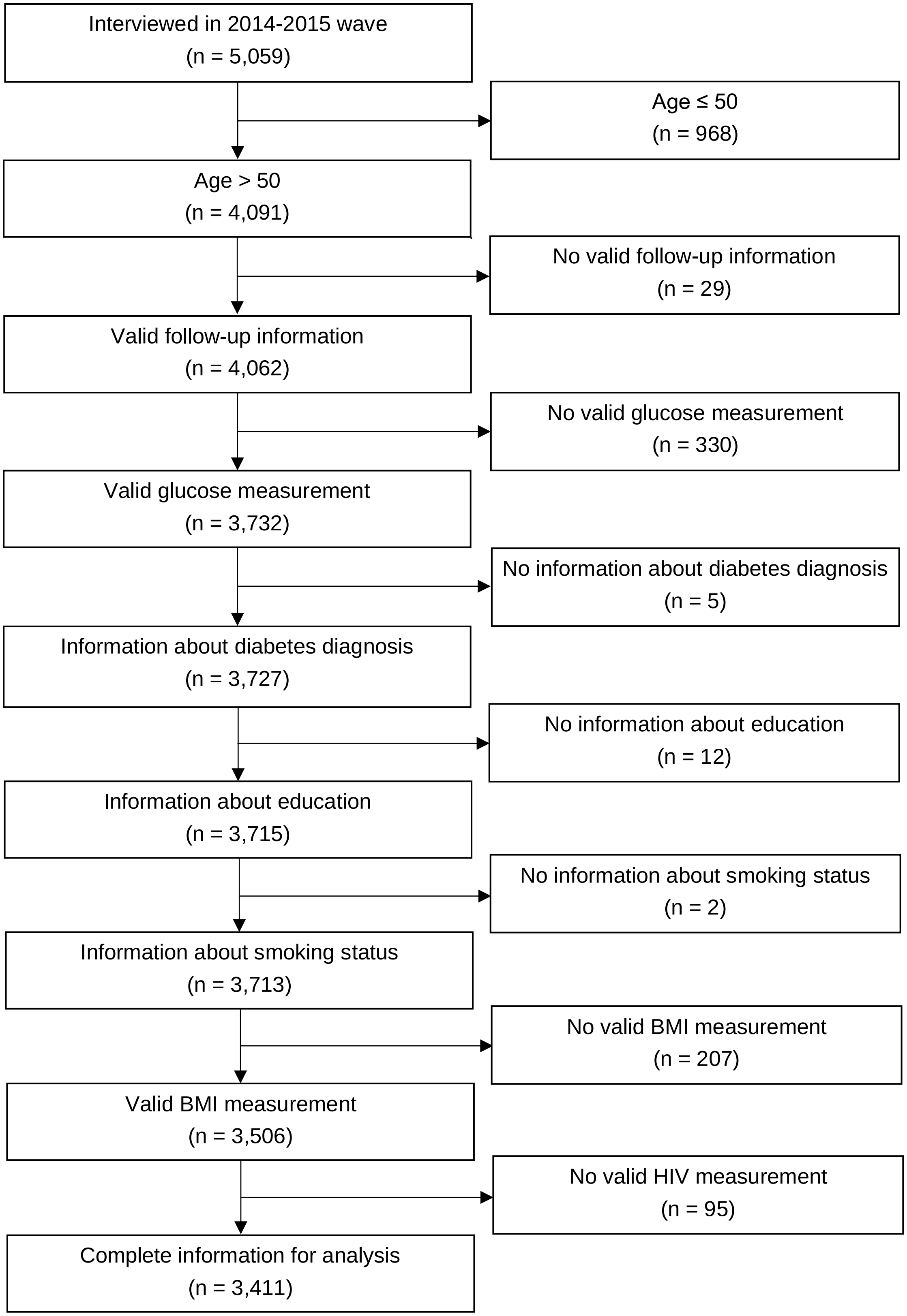

Health and Retirement Study (HRS)

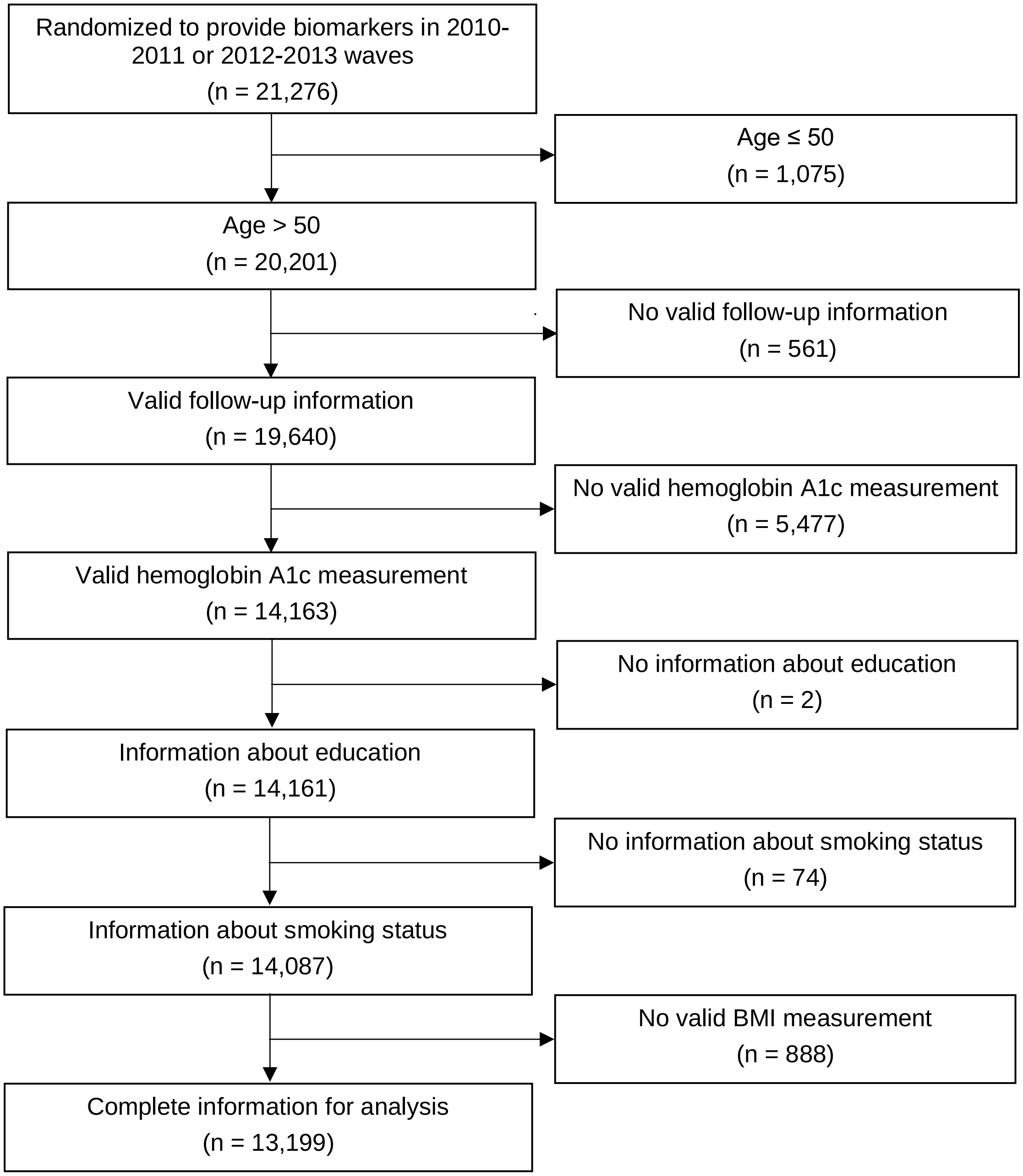

**Appendix 3: Text of relevant questions in the underlying cohorts**

| Study | Diabetes diagnosis question | Diabetes glucose-lowering medication question(s) |
|---|---|---|
| China (CHARLS) | Have you been diagnosed with diabetes or high blood sugar by a doctor? | Are you now taking any of the following treatments to treat or control your diabetes?<br><br>- Taking Western modern medicine<br>- Taking insulin injections |
| England (ELSA) | Has a doctor ever told you that you have diabetes? | Do you currently inject insulin for diabetes?<br><br>Are you currently taking any tablets, pills or other medication that you swallow for diabetes? |
| Mexico (MHAS) | Has a doctor or medical personnel ever diagnosed you with diabetes? | Are you currently taking any oral medication in order to control your diabetes?<br><br>Are you currently using insulin shots? |
| South Africa (HAALSI) | [Women] Have you ever been told by a doctor, nurse, or other healthcare worker that you have raised blood sugar or diabetes outside of pregnancy?<br><br>[Men} Have you ever been told by a doctor, nurse, or other healthcare worker that you have raised blood sugar or diabetes? | Are you currently receiving any treatment for diabetes prescribed by a doctor, nurse, or other healthcare worker? |
| United States (HRS) | Has a doctor ever told you that you have diabetes or high blood sugar? | In order to treat or control your diabetes, are you now taking medication that you swallow?<br><br>Are you now using insulin shots or a pump? |

**Appendix 4: Details on diabetes blood-based biomarkers used in each cohort**

| Study | Diabetes biomarker | Justification for biomarker selection | Collection and assay details |
|---|---|---|---|
| China (CHARLS)[1] | Glucose | HbA1c values are available, but study investigators raised concerns that HbA1c values may be underestimated due to potential degradation during transport and storage of frozen samples.[1] | Venous blood was collected by trained staff from the China Center for Disease Control and Prevention (CDC). Samples were separated into plasma and buffy coat and stored at -20 degrees Celsius immediately and during shipment, within two weeks, to the China CDC in Beijing for storage at -80 degrees Celsius. Samples were carried to the Youanmen Center for Clinical Laboratory of Capital Medical University, also in Beijing, for testing. Glucose was measured between February 2013 and June 2013 using an enzymatic colorimetric test. |
| England (ELSA)[2,3] | HbA1c | Fasting glucose values were only available for a subsample of participants fasting blood samples. Respondents over 80 years, known to be diabetic and on treatment, with a clotting or bleeding disorder or on anti-coagulant drugs, who had ever had fits, who seemed frail, or respondents, or whose health was a cause for concern were not asked to fast. | Venous blood was collected by qualified nurses. Samples were sent to the Royal Victoria Infirmary in Newcastle upon Tyne for testing. HbA1c was measured using the Tosoh G8 analyzer (Tosoh Bioscience, Tokyo, Japan). |
| Mexico (MHAS)[4] | HbA1c | Fasting glucose values are not available in the baseline wave used in this analysis. | Capillary blood was collected after a fingerstick. HbA1c was measured using the point-of-care A1CNow System (PTS Diagnostics, Whitestown, Indiana, USA), a method certified by the National Glycohemoglobin Standardization Program. |
| South Africa (HAALSI)[5] | Glucose | Dried blood hemoglobin A1c was collected, but plasma equivalent values are not available at present. | Capillary blood was collected after a fingerstick. Glucose was measured using the point-of-care CareSens N Monitor (i-SENS, Seoul, South Korea). |
| United States (HRS)[6] | HbA1c | Fasting glucose values are not available in the baseline wave used in this analysis. | Capillary blood was collected after a fingerstick using a dried blood spot (DBS) card. Samples that were collected during the 2010-2011 wave were first sent to the University |

| | | | |
|---|---|---|---|
| | | | of Michigan in Ann Arbor for storage and then to Heritage Laboratory in Olathe, Kansas for testing. HbA1c was measured using the Appraise test.<br>Samples that were collected during the 2012-2013 wave were first sent to the University of Michigan in Ann Arbor for storage and then to the University of Washington Department of Medicine Dried Blood Spot Laboratory in Seattle for testing. At the University of Washington, samples were stored at -80 degrees Celsius until the time of testing. From blood spots, punches of 3.2 mm in diameter were used to measure HbA1c using the Bio-Rad Variant II Hemoglobin Testing System (Bio-Rad Laboratories, Hercules, California, USA). Because clinical HbA1c cut-off points for diagnosis of diabetes are based on whole blood and not DBS values, this analysis used the National Health and Nutrition Examination Survey (NHANES) equivalent values that are provided by the HRS. Briefly, the distribution of the weighted HRS DBS values were adjusted to be consistent with the distribution of the weighted NHANES venous blood values. The 2010-2011 and 2012-2013 HRS samples were compared to the pooled 2009-2010 and 2011-2012 NHANES samples. |

**Appendix 5: Conceptual modeling using directed acyclic graph (DAG)**

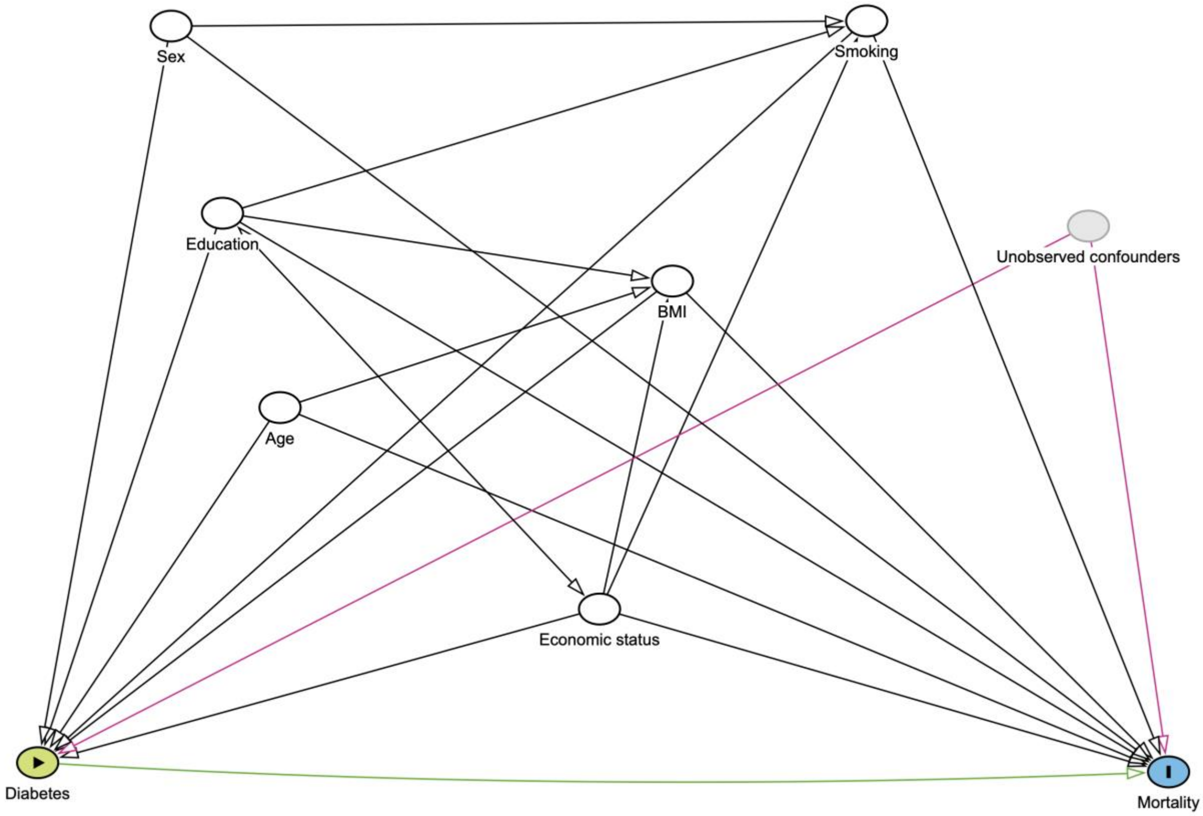

Examples of unobserved confounders include genetic ancestry, race/racism, diet, physical activity, occupation, other environmental exposures, and other variables. Note that HIV status is not shown here but is adjusted for in analyses in the South African cohort.

**Appendix 6: Adjusted all-cause mortality rates by cohort**

| | China (CHARLS) | England (ELSA) | Mexico (MHAS) | South Africa (HAALSI) | United States (HRS) |
|---|---|---|---|---|---|
| Mortality rate | | | | | |
| Without diabetes | 19.8 (17.7-21.9) | 17.0 (14.6-19.3) | 14.3 (9.3-19.3) | 32.9 (29.8-36.0) | 25.6 (23.4-27.8) |
| Diabetes | 35.5 (28.6-42.4) | 28.8 (22.1-35.6) | 29.0 (19.0-39.0) | 57.5 (45.5-69.5) | 39.2 (36.1-42.4) |

Mortality rates are presented as the number of deaths per 1,000 person years. Estimates were derived using Poisson regression models with an offset for log-transformed person-years and robust standard errors and adjusted for age, gender, education, smoking status, body mass index, and economic status. Models in South Africa also adjusted for HIV status. CHARLS=China Health and Retirement Longitudinal Study. ELSA=English Longitudinal Study of Ageing. HAALSI=Health and Aging in Africa: A Longitudinal Study of an INDEPTH Community in South Africa. HRS=Health and Retirement Study. MHAS=Mexican Health and Aging Study.

**Appendix 7: Mortality rate ratios by age groups**

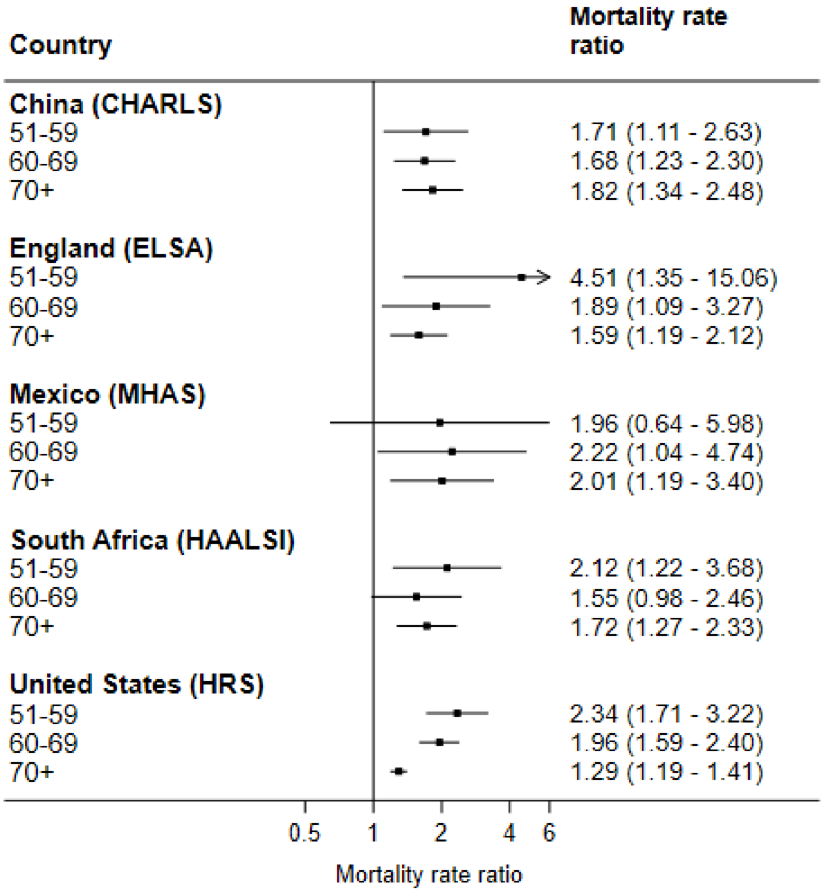

The horizontal error bars represent 95% CIs. Estimates were derived using Poisson regression models with an offset for log-transformed person-years and robust standard errors and adjusted for gender, education, smoking status, body mass index, and economic status. Models in South Africa also adjusted for HIV status. CHARLS=China Health and Retirement Longitudinal Study. ELSA=English Longitudinal Study of Ageing. HAALSI=Health and Aging in Africa: A Longitudinal Study of an INDEPTH Community in South Africa. HRS=Health and Retirement Study. MHAS=Mexican Health and Aging Study.

### Appendix 8: Comparisons of diagnosed vs. undiagnosed

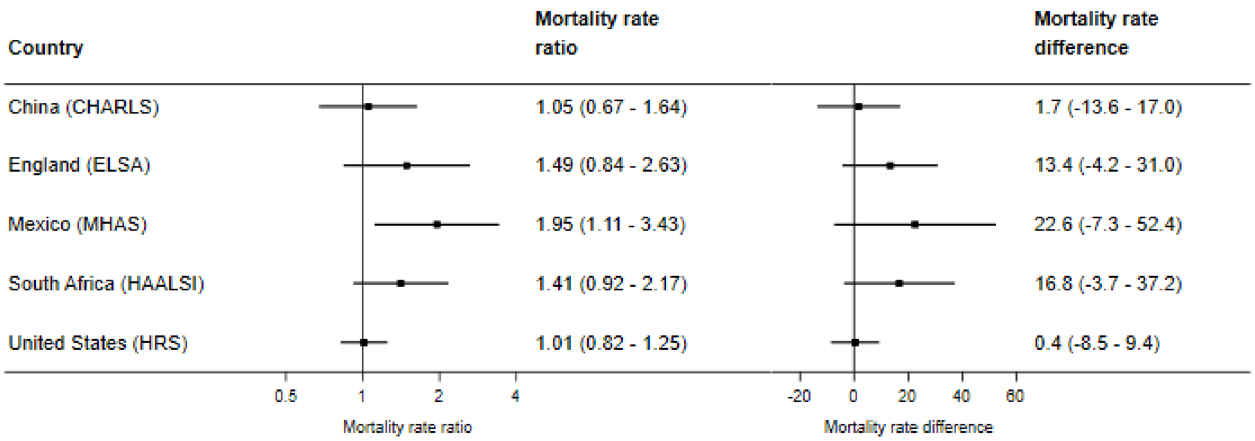

Mortality rate differences are presented as the number of deaths per 1,000 person years. The horizontal error bars represent 95% CIs. Estimates were derived using Poisson regression models with an offset for log-transformed person-years and robust standard errors and adjusted for age, gender, education, smoking status, body mass index, and economic status. Models in South Africa also adjusted for HIV status. CHARLS=China Health and Retirement Longitudinal Study. ELSA=English Longitudinal Study of Ageing. HAALSI=Health and Aging in Africa: A Longitudinal Study of an INDEPTH Community in South Africa. HRS=Health and Retirement Study. MHAS=Mexican Health and Aging Study.

**Appendix 9: Sensitivity analysis using Cox proportional hazards models instead of Poisson with an offset for log-transformed person-years and robust standard errors**

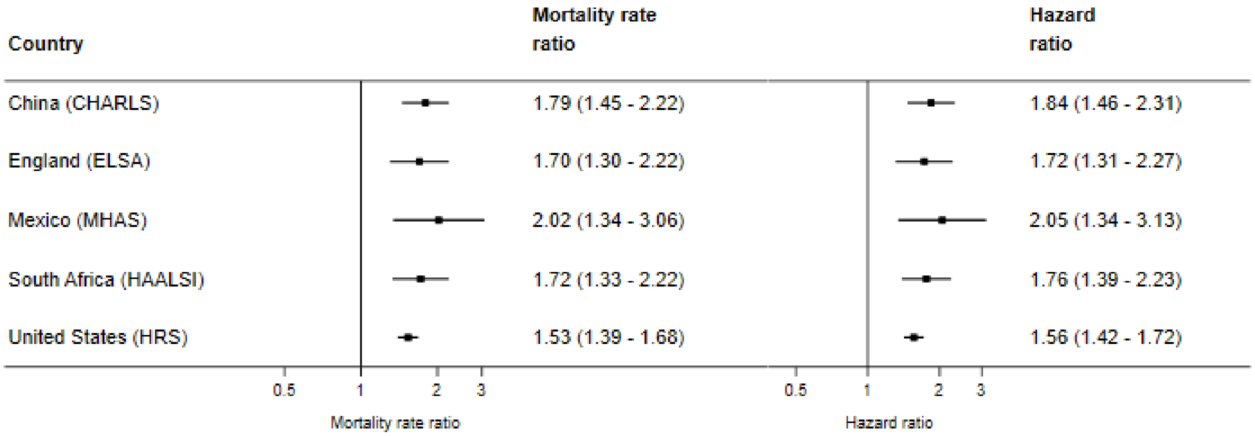

The horizontal error bars represent 95% CIs. Mortality rate ratios were derived using Poisson regression models with an offset for log-transformed person-years and robust standard errors. Hazard ratios were derived using Cox proportional hazards models. Models adjusted for age, gender, education, smoking status, body mass index, and economic status. Models in South Africa also adjusted for HIV status. CHARLS=China Health and Retirement Longitudinal Study. ELSA=English Longitudinal Study of Ageing. HAALSI=Health and Aging in Africa: A Longitudinal Study of an INDEPTH Community in South Africa. HRS=Health and Retirement Study. MHAS=Mexican Health and Aging Study.

### Appendix 10: Sensitivity analysis using self-report of diabetes medication instead of self-report of diabetes diagnosis

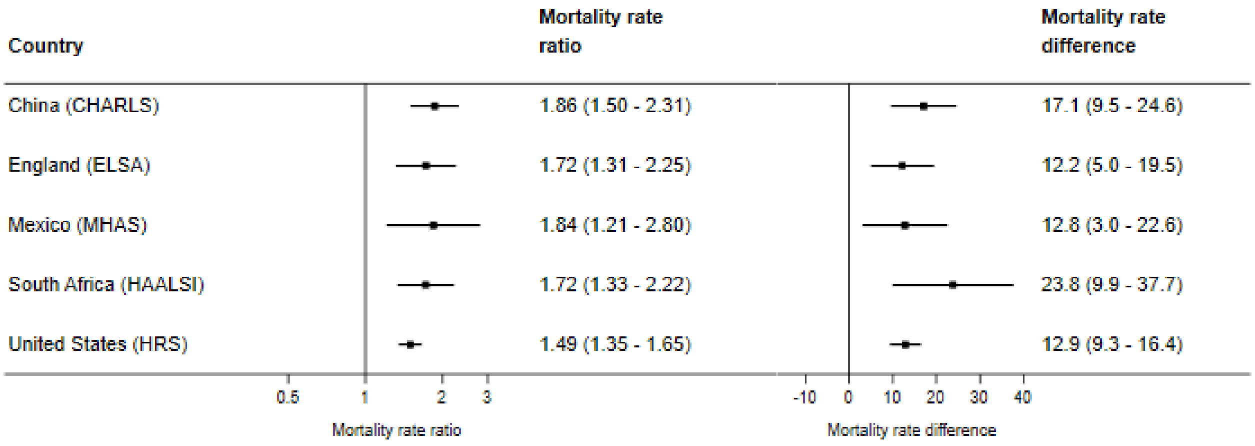

Mortality rate differences are presented as the number of deaths per 1,000 person years. The horizontal error bars represent 95% CIs. Estimates were derived using Poisson regression models with an offset for log-transformed person-years and robust standard errors and adjusted for age, gender, education, smoking status, body mass index, and economic status. Models in South Africa also adjusted for HIV status. CHARLS=China Health and Retirement Longitudinal Study. ELSA=English Longitudinal Study of Ageing. HAALSI=Health and Aging in Africa: A Longitudinal Study of an INDEPTH Community in South Africa. HRS=Health and Retirement Study. MHAS=Mexican Health and Aging Study.

### Appendix 11: Sensitivity analysis not adjusting for BMI

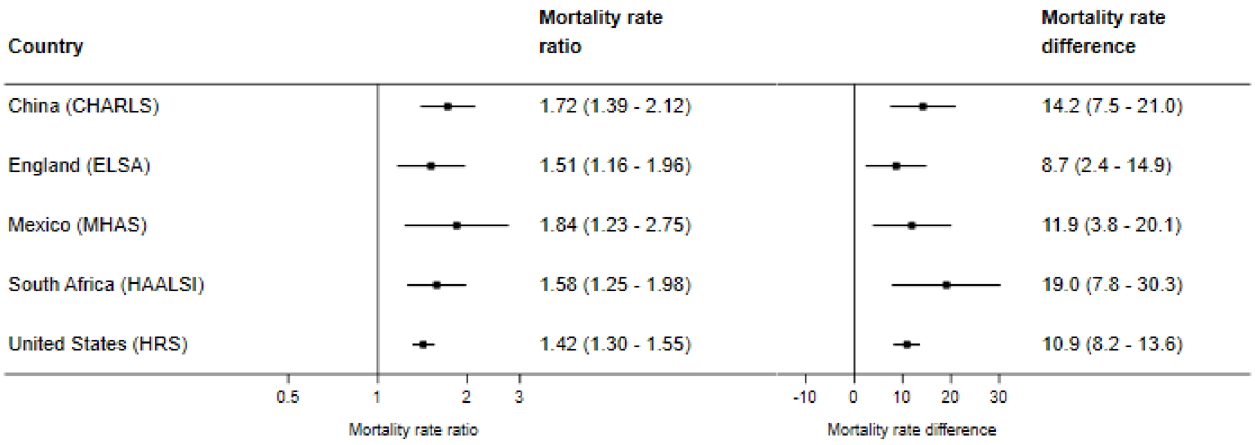

Mortality rate differences are presented as the number of deaths per 1,000 person years. The horizontal error bars represent 95% CIs. Estimates were derived using Poisson regression models with an offset for log-transformed person-years and robust standard errors and adjusted for age, gender, education, smoking status, and economic status. Models in South Africa also adjusted for HIV status. CHARLS=China Health and Retirement Longitudinal Study. ELSA=English Longitudinal Study of Ageing. HAALSI=Health and Aging in Africa: A Longitudinal Study of an INDEPTH Community in South Africa. HRS=Health and Retirement Study. MHAS=Mexican Health and Aging Study.
